# Common factor GWAS identifies shared risk loci between chronic widespread pain, atherosclerosis and arterial stiffness

**DOI:** 10.1101/2025.09.08.25331262

**Authors:** Dúalta Padraigh McGrath, Maryam Kazemi Naeini, Maxim B Freidin, Marina Cecelja, Roger Compte, Frances MK Williams

## Abstract

Chronic widespread pain (CWP) is a complex condition linked to impaired arterial health, including atherosclerosis and increased arterial stiffness. Epidemiological evidence suggests shared biological mechanisms, with strong associations between CWP and arterial dysfunction, However, the genetic basis remains largely unexplored.

We conducted a common pathway genome-wide association study using genomic structural equation modeling (GenomicSEM) and GWAS summary statistics for CWP, atherosclerosis, and pulse wave velocity to identify shared genetic factors.

This analysis revealed 53 genome-wide significant variants contributing to a shared latent factor, with opposing trait loadings suggestive of antagonistic pleiotropy. Lead loci included RNF123, ATP2C1, and COMT, with gene-level analysis implicating neurodevelopmental pathways and glycosaminoglycan degradation through hyaluronidase activity. Chromatin interaction and expression mapping supported regulatory links in relevant tissues.

Our findings demonstrate that neurogenic and extracellular matrix-related processes, including glycan metabolism, contribute to the shared genetic architecture of CWP and cardiovascular traits, offering mechanistic insight into their comorbidity.

## Introduction

Chronic widespread pain (CWP) is a prevalent musculoskeletal disorder, affecting 10–15% of adults globally, with a higher prevalence in women and older individuals^1^. The American College of Rheumatology (ACR) defines CWP as pain persisting for at least three months above and below the waist, on both sides of the body, and in the axial skeleton^2^. CWP often co-occurs with fatigue, sleep disturbance, and diminished self-esteem^3^, anxiety and depression^4^, exacerbating disability, work limitations, and increased healthcare costs.

Genetic studies demonstrate the heritability of CWP, with Burri et al.^5^ estimating CWP heritability at 58%, consistent with previous estimates of 46–54%^6–8^. Genome-wide association studies (GWAS) have further characterised CWP genetics. Rahman et al.^9^ estimated SNP-based heritability at 33% in a UK Biobank Northern European population, identifyingseveral loci contributing to CWP risk, including *RNF123*.

Despite its societal impact, CWP’s molecular mechanisms remain unclear, limiting therapeutic development. Small-sample clinical studies have suggested that variants involved in immune response and inflammation may contribute to the development of CWP. Generaal et al.^10^, using a sample of 1,632 individuals, reported inflammation and immune activation in musculoskeletal pain, though larger studies have not consistently supported systemic inflammation, despite possible immune pathway involvement in CWP risk.

CWP commonly co-occurs with cardiovascular disease. Macfarlane et al.^11^ reported a strong association between CWP and excess cardiovascular-related mortality, independent of confounding factors like age, BMI, and smoking, highlighting the need to identify shared biological and genetic risk factors contributing to this comorbidity.

Building on this, Naeini et al.^12^ investigated genetic correlations between CWP and cardiovascular traits, identifying a possible latent factor underlying CWP, atherosclerosis, and arterial stiffness using twin models. This latent factor captures shared SNP-based variance across multiple traits, while accounting for pleiotropy and trait-specific effects. In their model, genetic factors explained 68% of latent factor variation in carotid-femoral pulse wave velocity (cfPWV) and 90% in carotid plaques. This reflects a broader principle in complex disorders like CWP, where genes often display pleiotropic effects^13^, influencing multiple phenotypic outcomes and the pathology of other conditions.

Glycans, including glycosaminoglycans, are key components of extracellular matrix remodelling, tissue integrity, inflammation, and pain signalling. Altered glycan metabolism may link neuroinflammation and vascular dysfunction, potentially explaining the overlap between chronic pain and cardiovascular outcomes. Disrupted glycosaminoglycan homeostasis has been implicated in cartilage degradation, joint pain, vascular stiffening, and plaque formation, reinforcing its relevance to both phenotypes^14^.

The identification of a shared latent factor linking CWP, atherosclerosis, and arterial stiffness aligns with pleiotropy in complex diseases^13^, where shared single nucleotide polymorphisms (SNPs) contribute to multiple disorders. This suggests common pathophysiological mechanisms, with genes potentially influencing immune regulation, vascular function, and pain signalling simultaneously.

Arterial stiffness is a key intermediate phenotype linking CWP to atherosclerosis, measured by cfPWV. It results from elastin and collagen alterations in arterial walls, contributing to atherosclerotic plaque^15^. It is a strong cardiovascular risk indicator^16^ and may therefore serve as a biomarker in CWP.

Atherosclerosis is characterised by arterial atheroma formation. As it progresses, arterial stiffness and narrowing increase heart attacks and stroke risk, particularly when mature atheromas rupture, causing local clotting and vessel occlusion^17^.

Chronic inflammation drives atherosclerosis^18^. Inflammatory cytokines may link CWP and atherosclerosis, as both conditions may involve common inflammatory pathways. Shared genetic variation may also contribute to the overlapping pathophysiological mechanisms between CWP and atherosclerosis.

Advances in GWAS, particularly larger meta-analyses, have improved variant detection in complex traits¹⁹ and enabled multivariate approaches like Genomic Structural Equation Modelling (GenomicSEM) to uncover shared genetic architecture across disorders.

The utility of GenomicSEM is exemplified by Zorina-Lichtenwalter et al.^20^, who identified a general genetic factor underlying 24 chronic pain conditions. GenomicSEM distinguishes between general shared pathways and condition-specific genetic factors. Additionally, the tool confirmed previously discovered novel SNPs, highlighting its potential to advance our understanding of the shared genetic architecture of complex disorders.

Our study builds on Naeini et al., who used twin data and modelling to find shared genetic and environmental factors driving arterial stiffness, carotid plaque formation, and CWP. We aimed to identify genetic variants that contribute to the common latent component shared by atherosclerosis, arterial stiffness, and CWP. We applied a common factor model to GWAS datasets for arterial stiffness, atherosclerosis, and CWP to gain new understanding of the common pathways that may raise cardiovascular risk in people with CWP.

## Results

### GWAS Datasets and SNP Filtering

GWAS summary statistics were processed for GenomicSEM common factor GWAS. Key dataset characteristics, including sample size and SNP retention, are detailed in Supplementary Table 1. The CWP dataset included 6,914 cases and 242,929 controls, while the atherosclerosis dataset included 14,334 cases and 346,860 controls. Arterial stiffness was analysed as a continuous trait in 151,053 individuals.

Each trait’s dataset initially included millions of SNPs. After filtering to include well-imputed HapMap3 SNPs for LDSC matrices, each trait retained over one million SNPs. A final set of 5,952,753 SNPs shared across datasets was included in the model, ensuring an overlapping and consistent variant set across traits.

### Trait Heritability Estimates

All traits showed significant SNP-based heritability (see Supplementary Table 2). Heritability was higher for CWP and atherosclerosis compared to arterial stiffness, aligning with previously reported estimates^6–8^.

### Genetic Correlations Between Traits

Pairwise genetic correlations were calculated using bivariate LDSC from HapMap3-filtered SNPs. CWP and atherosclerosis displayed a moderately strong negative correlation (−0.3446, SE = 0.0465). CWP and arterial stiffness also displayed a negative correlation (−0.1807, SE = 0.0608). In contrast, atherosclerosis and arterial stiffness showed a positive correlation (0.1817, SE = 0.0634). All estimates reached statistical significance (*P* < 0.005). These relationships are visualised in Figure 1.

**Figure 1:**
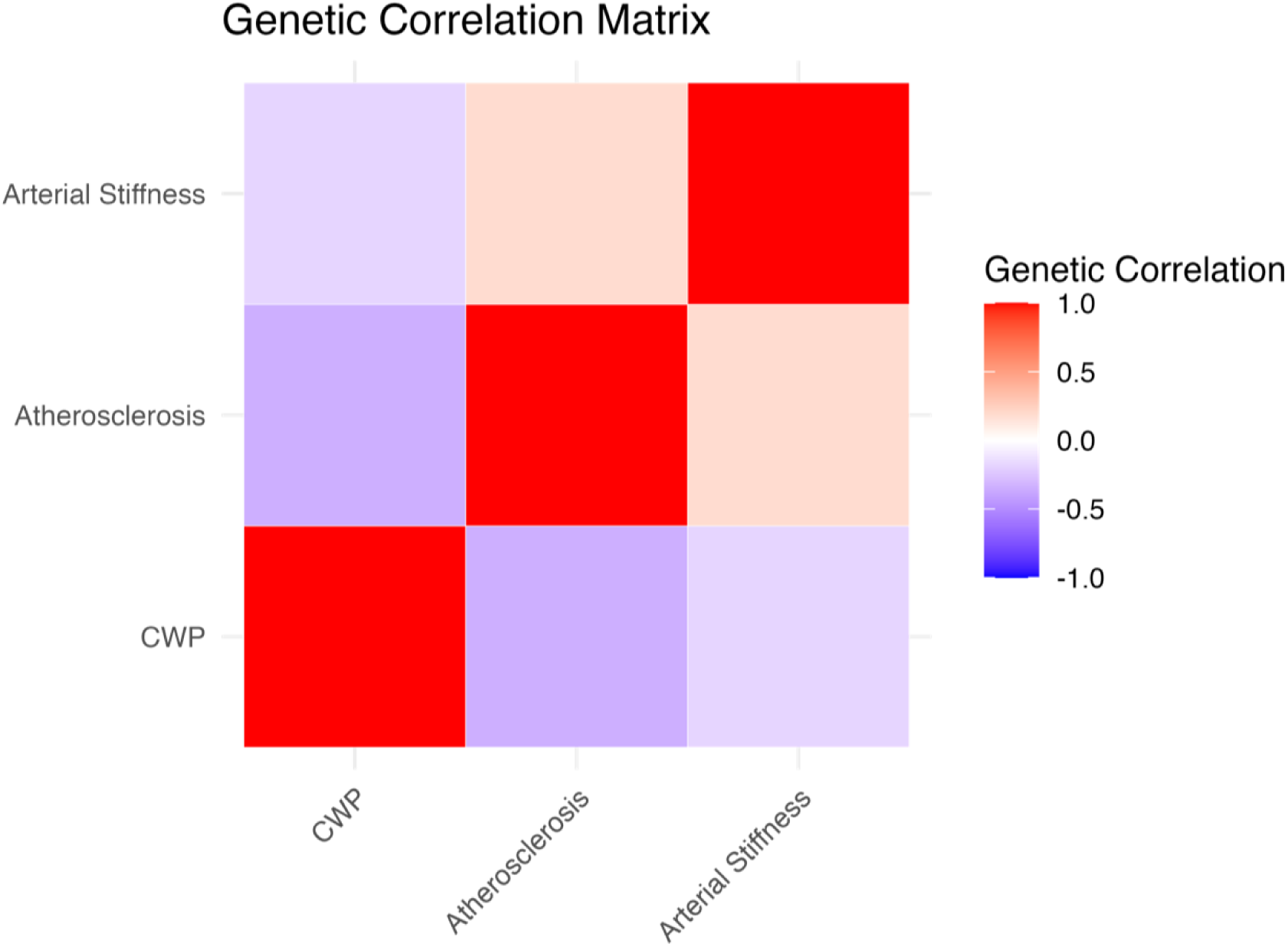
Heatmap of Genetic Correlation between Traits. Heatmap visualising genetic correlations between traits. The plot was generated using LDSC-derived correlation estimates Colours represent the strength and direction of the correlations, with red indicating positive genetic correlations and blue indicating negative genetic correlations. The heatmap was created in R using ggplot2.

### Common Factor Model Loadings

Standardised factor loadings from the common factor model are presented in Figure 2. The loading for CWP was fixed at 1.0, while the loadings for atherosclerosis and arterial stiffness were estimated from summary statistics,

**Figure 2:**
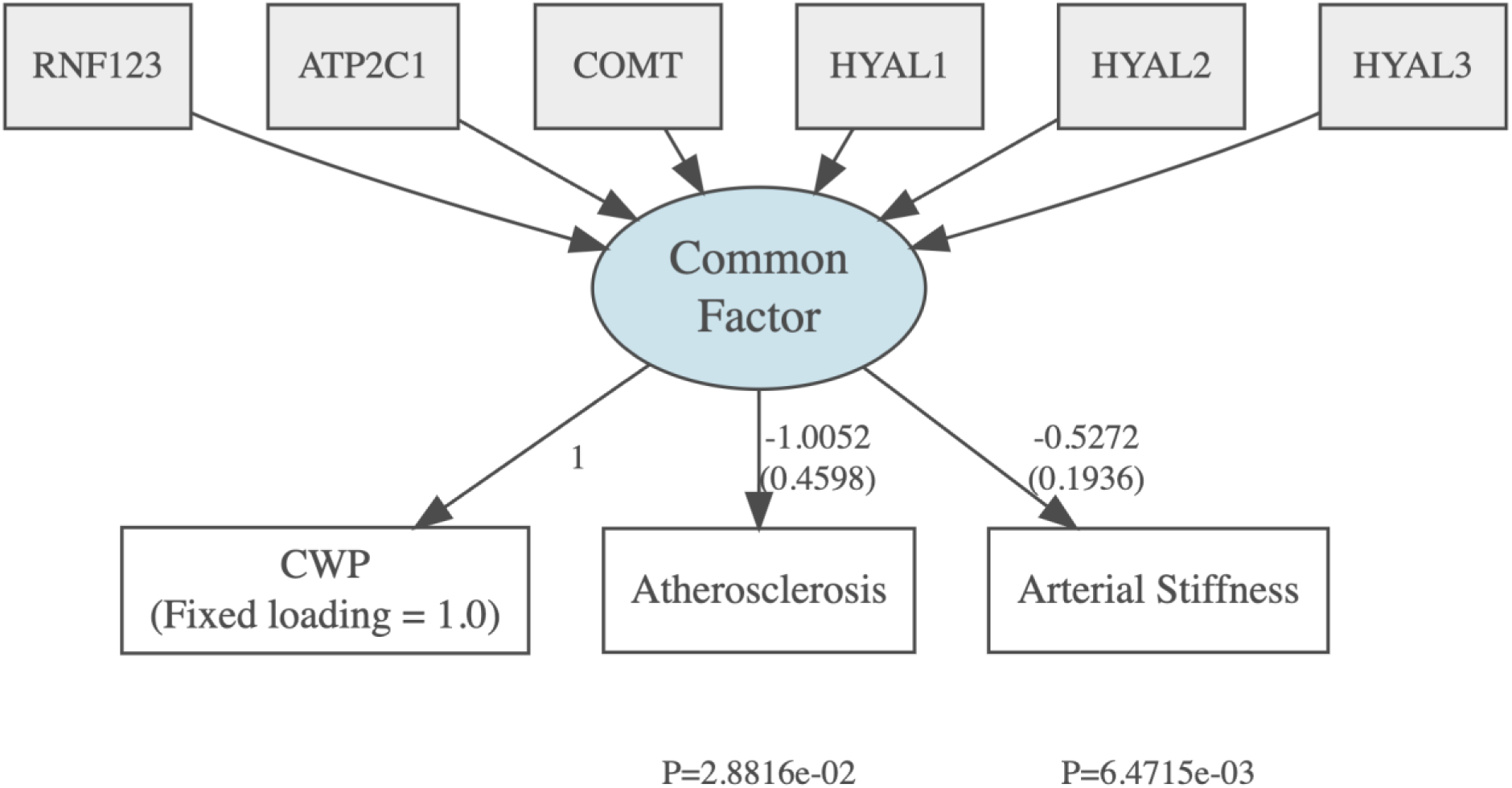
Standardised Factor Loadings from Common Factor GWAS Model. Standardised factor loadings from the common factor model. CWP loading was fixed at 1.0 to define the latent variable scale, while atherosclerosis and arterial stiffness loadings were estimated based on their genetic covariance with CWP, with decimal values rounded to four decimal places. Standard error (SE) values are indicated in brackets below factor loadings, and P indicates the P-value for each trait’s estimated loading. Six gene loci (RNF123, ATP2C1, COMT, HYAL1, HYAL2, HYAL3) are shown contributing to the latent factor within the model. Genes included in the diagram were identified through gene mapping analyses described in subsequent sections.

Atherosclerosis and arterial stiffness loaded negatively onto the latent factor (−1.0052 and –0.5272, respectively) suggesting inverse relationships with CWP. These findings indicate overlapping genetic influences across the traits. Atherosclerosis showed stronger negative loading with greater uncertainty (SE = 0.4598), possibly indicating greater variability in its genetic divergence from CWP, while arterial stiffness had a smaller negative loading with more precise estimation (SE = 0.1936).

### Common Factor GWAS Output Summary

A common factor GWAS was performed across the 22 autosomal chromosomes to identify genetic variants contributing to shared liability across CWP, atherosclerosis, and arterial stiffness. Sex chromosomes were excluded due to differing inheritance patterns requiring separate analysis beyond the scope of this study.

Latent factor SNP-based heritability was estimated at 3.44% (SE = 0.0028) using LDSC, with a Z-score of 12.3 (*P <* 1e-30) indicating strong statistical significance.

SNPs were evaluated for suggestive (*P* < 5e-6) and genome-wide (*P* < 5e-8) significance. To distinguish SNPs contributing to the latent factor from those driven by trait-specific heterogeneity, Q P-value filtering was applied. A high Q P-value (*Q P* > 0.05) represents homogeneous SNP effects, well explained by the latent factor, while low Q P-values (*Q P* < 0.05) reveal residual heterogeneity, where SNPs may act through additional trait-specific pathways.

In total, 590 SNPs exceeded suggestive significance, and 78 surpassed genome-wide significance. After Q P-value filtering, 424 and 53 SNPs remained, respectively, indicating that many SNP associations may reflect trait-specific rather than shared genetic effects. This filtering approach prioritised SNPs likely to reflect genuine shared genetic liability while reducing potential false positives from heterogeneity. SNP counts and thresholds are detailed in Supplementary Table 3. A Q-Q plot confirming model calibration and SNP enrichment is provided in Supplementary Figure 1.

### Genome-wide association signals

A Manhattan plot visualising the GWAS results is shown in Figure 3, displaying −log₁₀(P) values for 5,952,753 SNPs tested for association with the latent factor across all autosomes. Genome-wide (P < 5e-8) and suggestive (P < 5e-6) significance thresholds are shown with red and blue horizontal lines, respectively. SNPs passing genome-wide significance and Q P-value filtering were annotated with gene labels, prioritising the most significant SNP per gene.

**Figure 3:**
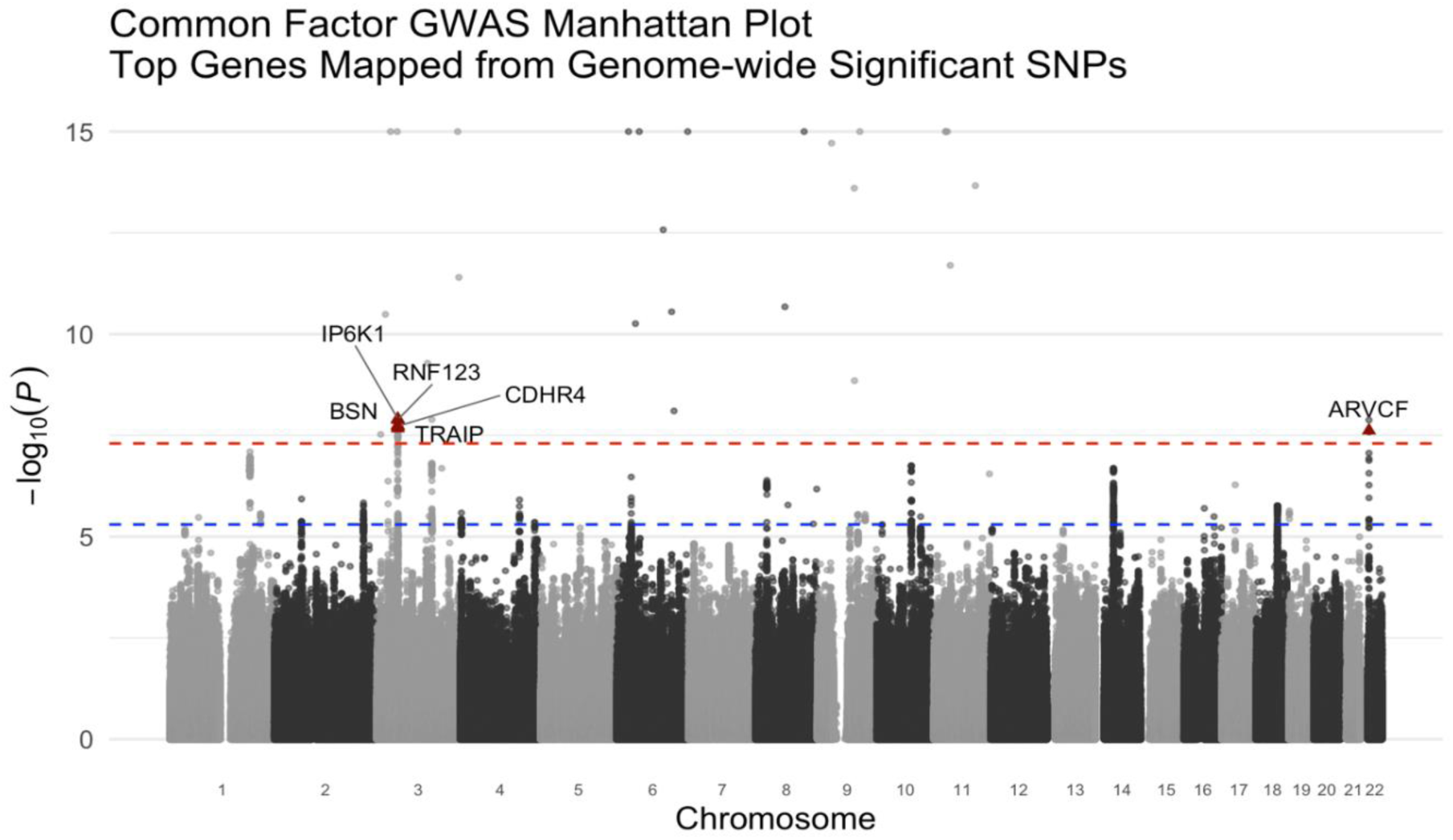
Manhattan Plot of the Common Factor GWAS. Manhattan plot showing genome-wide association results from the common factor GWAS created using ggplot2 in R. Each point represents a SNP tested for association with the latent factor. The x-axis shows the genomic position by chromosome and the y-axis shows −log₁₀(P) values for association with the latent factor, capped at 15 for visual clarity. Genome-wide (P < 5e-8) and suggestive significance (P < 5e-6) thresholds are indicated by red and blue horizontal lines, respectively. Genome-wide significant SNPs passing Q P-value filtering were annotated with gene labels. Genes labels were applied once, prioritising the most significant variant per gene. These represent the most significant SNPs mapping to genes flagged multiple times in the filtered dataset (Table 3).

**Table 3:** Genome-wide significant SNPs associated with the Latent Factor. Genome-wide significant SNPs identified from the common factor GWAS. All SNPs passed Q P-value filtering (Q P > 0.05), suggesting their effects are well explained by the latent factor. Gene symbols and functional consequences were obtained using the Open Targets Platform. Where a SNP is mapped to multiple genes, each gene is shown in a separate row. Where multiple SNPs mapped to a gene, only the most significant SNP (lowest P-value) was retained. Effect size estimates reflect the beta coefficients representing each SNPs association with the latent factor. Gene recurrence indicates the number of times a SNP is mapped to each gene across the full filtered set of SNPs. All decimal values were rounded to four decimal places.

| SNP (rsID) | Gene | CHR | BP | P-value | Effect size estimate | Variant type | Gene recurrence |
| --- | --- | --- | --- | --- | --- | --- | --- |
| rs1491985 | RNF123 | 3 | 49754970 | 1.1906E-08 | +0.001920 | Intronic | 6 |
| rs7628207 | AMIGO3 | 3 | 49754970 | 1.2120E-08 | +0.001919 | 3' UTR | 1 |
| rs7628207 | GMPPB | 3 | 49754970 | 1.2120E-08 | +0.001919 | Downstream | 1 |
| rs9870858 | IP6K1 | 3 | 49769071 | 1.2297E-08 | +0.001917 | Intronic | 10 |
| rs10490825 | ATP2C1 | 3 | 130696383 | 1.2879E-08 | -0.002143 | Intronic | 1 |
| rs165599 | ARVCF | 22 | 19956781 | 1.3201E-08 | -0.001585 | 3' UTR | 3 |
| rs165599 | COMT | 22 | 19956781 | 1.3208E-08 | -0.001585 | 3' UTR | 1 |
| rs1352889 | BSN | 3 | 49652148 | 1.7763E-08 | +0.001898 | Intronic | 21 |
| rs7637711 | CDHR4 | 3 | 49829653 | 1.8739E-08 | -0.001895 | Intronic | 4 |
| rs62260723 | TRAIP | 3 | 49872124 | 2.1189E-08 | -0.001889 | Intronic | 4 |
| rs3020779 | MST1 | 3 | 49724808 | 2.4120E-08 | +0.001880 | Synonymous | 1 |
| rs4855882 | APEH | 3 | 49715354 | 2.5374E-08 | +0.001879 | Intron | 1 |
| rs62260720 | UBA7 | 3 | 49853796 | 3.2734E-08 | -0.001862 | Upstream | 2 |

A strong and densely clustered signal was observed on chromosome 3, including RNF123, IP6K1, BSN, CDHR4, and TRAIP. An additional genome-wide signal was detected on chromosome 22 at ARVCF.

Table 3 displays genome-wide significant SNPs that passed Q P-value filtering, indicating effects well explained by the latent factor. Gene annotations were derived from the Open Targets platform by selecting the gene with the highest variant-to-gene score per SNP. For SNPs mapping to multiple genes, each gene is listed separately, while for multiple SNPs mapping to the same gene, only the most significant variant was retained to highlight the top signal per locus. Gene recurrence reflects the number of SNPs mapping to each gene, indicating consistent association with the latent factor.

These results were drawn from 53 SNPs that exceeded genome-wide significance and passed Q P-value filtering, resulting in 13 unique gene entries. The most significant SNP, *rs1491985*, mapped to *RNF123* and was highlighted by six SNPs. Other notable entries included *rs9870858*, mapped to *IP6K1*, and *rs1352889,* mapped to *BSN*. Additional recurrent loci included *CDHR4, TRAIP*, and *ARVCF*.

While many variants were intronic, 3’ UTR, downstream, and synonymous variants were also observed, suggesting diverse mechanisms of shared genetic effects. Effect size directionality supported antagonistic pleiotropy, with several SNPs (e.g. *rs1491985*, *rs9870858)* associated with increased risk for CWP and reduced cardiovascular liability, while others (e.g. *rs10490825*, *rs165599*) showed the opposite pattern.

GO analysis revealed enrichment in neurodevelopmental processes, including synapse organisation and neuron development (see Supplementary Figure 2)

### Gene-based and prioritised variant analysis

To further prioritise SNPs, functional mapping and annotation were performed using FUMA to characterise the top loci and identify relevant tissues and molecular functions^21^.

FUMA identified 95 SNPs with three reaching independent significance (see Supplementary Table 4). These loci on chromosomes 3 and 22 included *RNF123, ATP2C1,* and *COMT,* also identified among the genome-wide significant SNPs in Table 3, reinforcing their relevance.

Gene-level association testing with MAGMA highlighted additional genome-wide hits (see Supplementary Figure 3), including BSN, CDHR4, TRAIP, RNF123, IP6K1, and UBA7 from Table 3. Additionally, *DPYSL2* and *MAML3,* not identified in the single SNP visualisation, emerged as novel signals.

### Tissue Enrichment and Differential Expression

MAGMA tissue expression analysis (Figure 4a) investigated whether the gene-level associations from the common factor GWAS aligned with gene expression levels across 54 tissues, quantified by transcripts per million (TPM). The strongest association signals appeared in brain regions, including the cerebellum, frontal cortex, and hippocampus, although none surpassed the Bonferroni-corrected threshold (P < 0.05/54). To further investigate tissue-specific expression, differentially expressed gene (DEG) analysis was performed using FUMA’s gene2func tool on the 95 prioritised genes mapped from suggestive SNPs.

**Figure 4:**
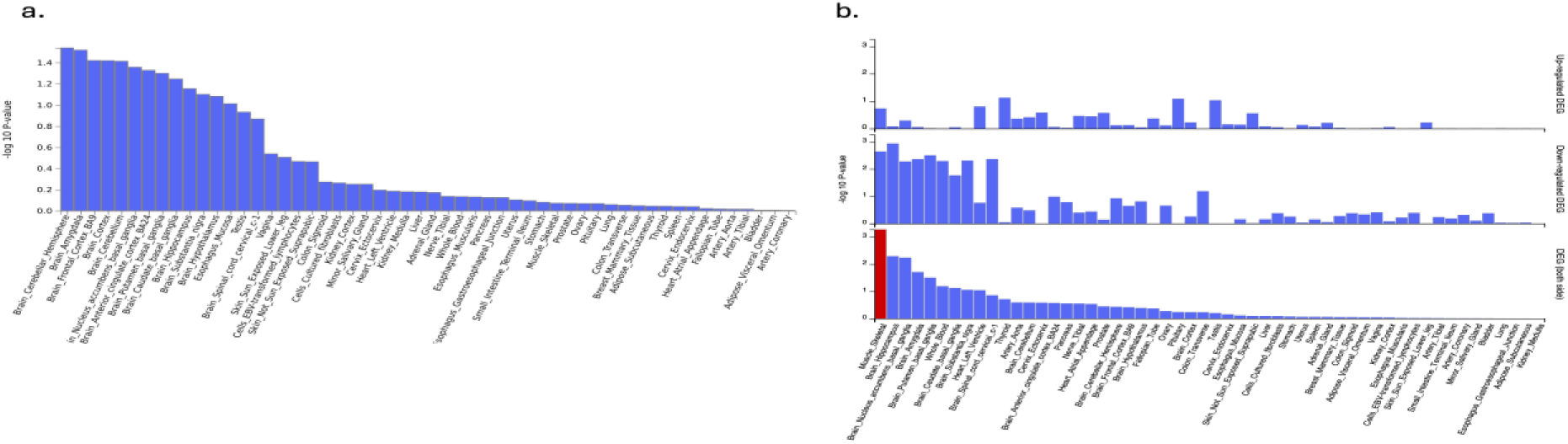
MAGMA Tissue Expression and Differential Expression of Prioritised Gene Results from GTEx v8. a: Tissue expression analysis using MAGMA to test whether gene-level associations were predicted by gene expression levels in 54 GTEx v8 tissues. Gene expression levels were measured using TPM. Tissues are labelled along the x-axis, while the y-axis represents the −log₁₀(P) values. No tissues surpassed Bonferroni-corrected significance (P < 0.05/54). b: Tissue-specific enrichment for DEGs across 54 GTEx v8 tissues based on the 95 genes mapped from suggestive SNPs using FUMA’s gene2func tool. Tissues are labelled along the x-axis, while the y-axis represents the −log₁₀(P) values. The three panels show upregulated DEGs (top), downregulated DEGs (middle), and DEGs in either direction (bottom). Tissues with significantly enriched DEGs are highlighted in red (P < 0.05/54).

Skeletal muscle was the only tissue to reach significance (P < 0.05/54) in the DEG analysis (Figure 4b). Several central nervous system (CNS) related tissues, including the hippocampus, amygdala, and spinal cord, showed consistent downregulation but did not pass the multiple testing threshold. MAGMA and DEG analyses support the interpretation of a predominantly nervous system-based latent factor, with additional relevance for muscle-specific biological mechanisms.

### Molecular Function and Pathway Enrichment

GO molecular function (GOMF) analysis (Figure 5a) identified overrepresented molecular mechanisms contributing to the latent factor. KEGG pathway enrichment (Figure 5b) highlighted broader biological pathways underlying the shared genetic liabiity.

**Figure 5:**
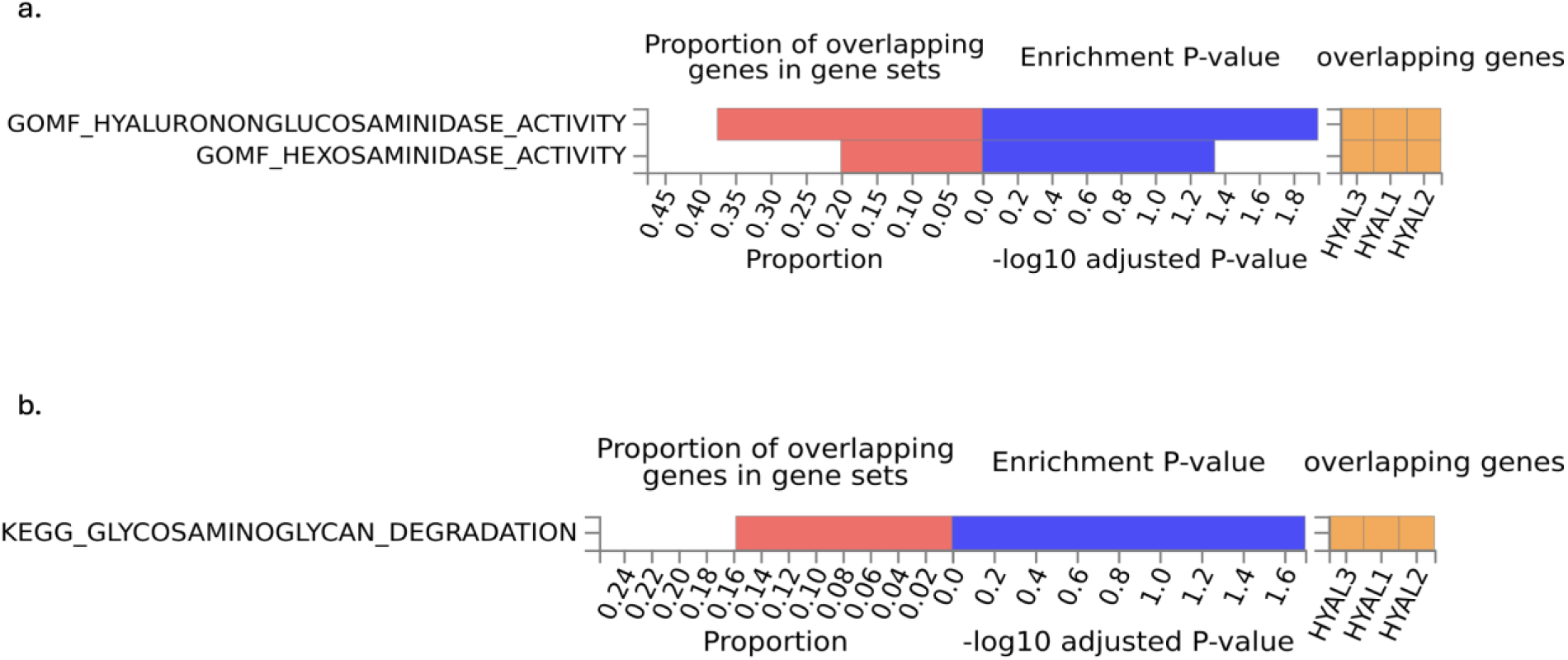
Functional Enrichment of Genes Contributing to the Latent Factor. a: GOMF enrichment of genes mapped from 424 SNPs with P < 5e-6 and Q P > 0.05. Top terms included synapse organisation, neuron development, and cell junction organisation. b: KEGG pathway enrichment using the same gene set, highlighting glycosaminoglycan degradation. Bars represent –log₁₀(P) enrichment values. Enrichment was assessed using g:Profiler with default gSCS multiple testing correction. Results implicate neurodevelopment and glycan metabolism in the shared genetic liability.

Enrichments were driven by *HYAL1*, *HYAL2*, and *HYAL3,* involved in hyaluronan degradation, a central process in extracellular matrix (ECM) remodelling and immune signalling. Hyalurononglucosaminidase and hexosaminidase enzymes contribute to the breakdown of glycosaminoglycans. While their activity is normal, excessive degradation can contribute to cartilage degeneration and joint dysfunction. Overlap between GO and KEGG suggests that glycan metabolism, particularly hyaluronan degradation, may underlie the latent factor.

To explore tissue specificity, Figure 6 presents expression profiles of *RNF123, ATP2C1, COMT, HYAL1*, *HYAL2*, and *HYAL3* across 54 tissue types.

**Figure 6:**
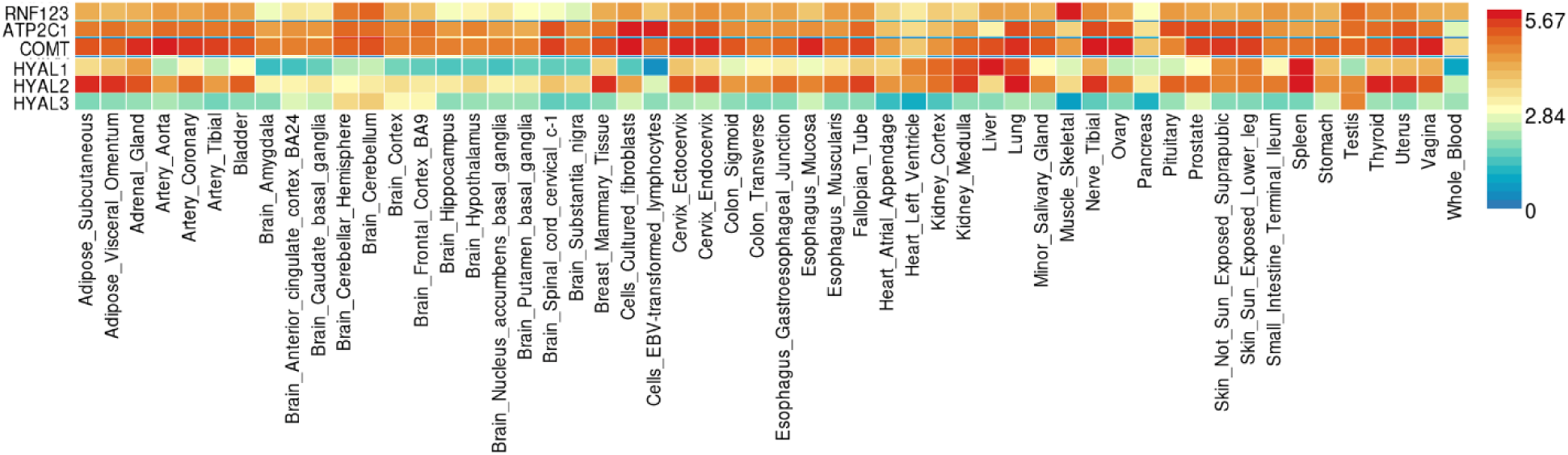
Tissue-specific expression of RNF123, ATP2C1, COMT, HYAL1, HYAL2, and HYAL3 across GTEx v8 tissues. Tissue-specific expression of RNF123, ATP2C1, COMT, HYAL1, HYAL2, and HYAL3 across 54 GTEx v8 tissues, measured in transcripts per million. The blue-to-red gradient indicates increasing gene expression levels (0 = blue, ≥ 5.67 = red).

*RNF123, ATP2C1,* and *COMT* displayed high expression across many tissues. *ATP2C1* and *COMT* showed strong expression in brain, vascular, and adipose tissues, while *RNF123* showed high skeletal muscle expression with lower levels in several brain regions

Among the *HYAL* genes, *HYAL2* exhibited the highest overall expression with strong adipose and arterial signals, and moderate levels in brain and muscle. *HYAL1* showed moderate expression in adipose tissue, but lower levels in brain and arterial tissues. *HYAL3* showed generally low expression across all tissues, with moderate expression in the cerebellum and frontal cortex. These expression patterns suggest that each *HYAL* gene may have unique roles in the biological processes underlying the latent factor.

### Chromatin Interaction and eQTL Mapping of Genomic Risk Loci

Chromatin interaction and eQTL mapping investigated regulatory effects of the three independent SNPs by linking them to genes using genome structure and expression data to predict relevant targets. Circos plots were generated for chromosomes 3 and 22 to visualise the regulatory landscape (Figure 7).

**Figure 7:**
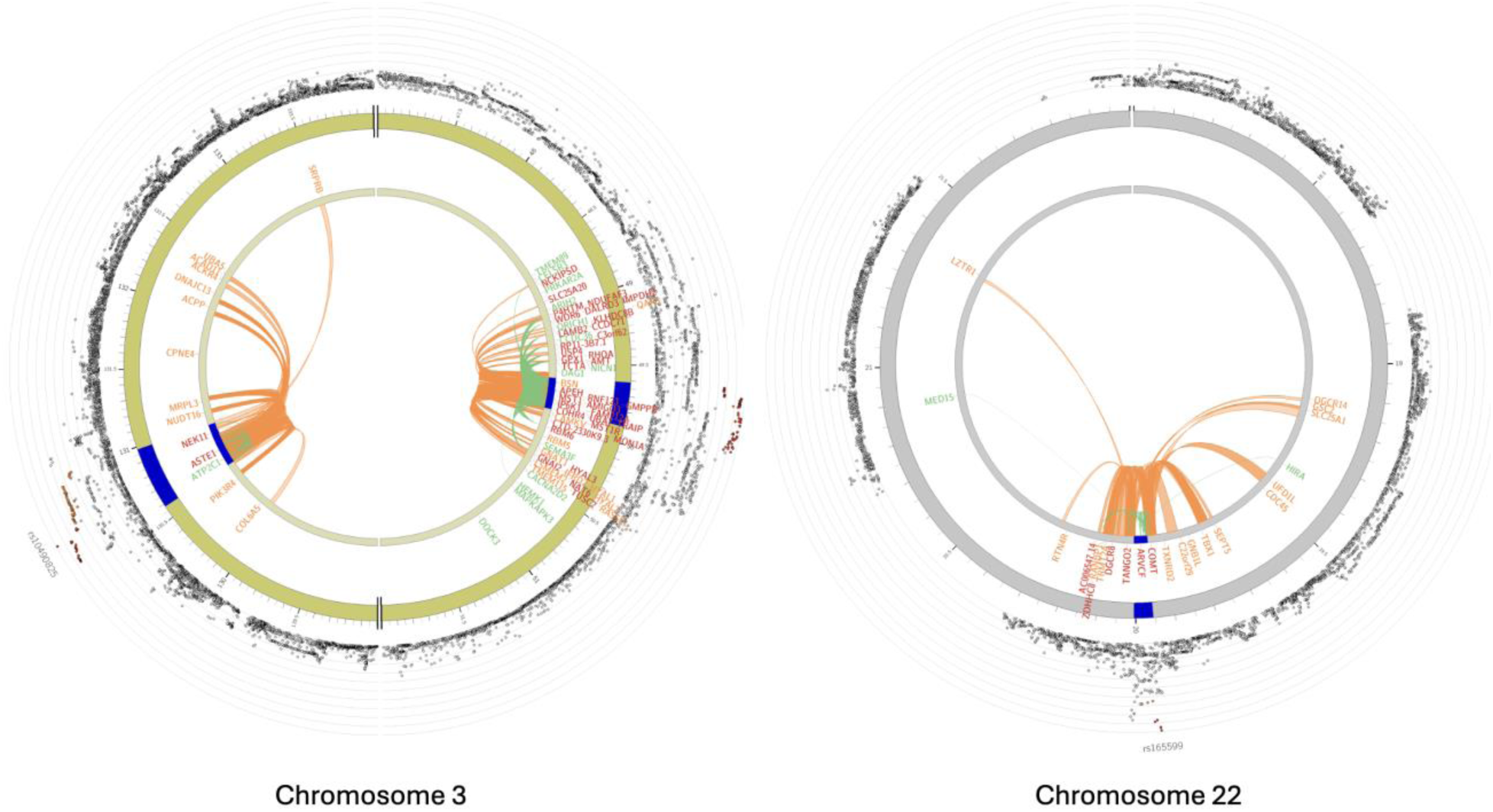
Chromatin Interaction and eQTL Mapping of Lead SNPs on Chromosome 3 and 22. Circos plots showing chromatin interaction and eQTL mapping of the genomic regions surrounding lead SNPs rs10490825 and rs1491985 (chromosome 3, left), and rs165599 (chromosome 22, right). The outermost layer is a Manhattan plot showing SNPs with P < 0.05, colour-coded by LD with lead SNPs which is measured by r² (squared correlation of allele frequencies): red (r² > 0.8), orange (r² > 0.6), green (r² > 0.4) and blue (r² > 0.2), and grey (r² ≤ 0.2) representing SNPs not in LD with lead SNPs. The second and third layers represent chromosomal coordinates, with risk loci highlighted in blue. Mapped genes are displayed on the inner layer, colour-coded by functional mapping: green (eQTL only), orange (chromatin interactions only), and red (both). Inner arcs represent functional connections: green represents eQTL links, while orange represents chromatin interaction links.

Candidate genes potentially regulated by the lead SNPs were identified. *HYAL1* and *HYAL2* were mapped through chromatin interactions only, while *HYAL3* was supported by chromatin interaction and eQTL mapping. Other recurrent genes, including *IP6K1* and *CDHR4* also appeared, reinforcing their potential contribution to the latent factor.

## Discussion

CWP is associated with increased cardiovascular risk, including atherosclerosis and arterial stiffness^11^. Despite epidemiological links, the genetic mechanisms underlying this comorbidity remain unclear. This study investigated the shared genetic architecture across these traits, hypothesising that immune and inflammatory-related SNPs would underlie their comorbidity. Analysis of approximately 5.95 million SNPs characterised the latent factor from Naeini et al.^12^, supported by Q-Q plot enrichment (see Supplementary Figure 1), indicating a polygenic signal and well-calibrated model.

Genetic correlations revealed CWP’s inverse relationship with atherosclerosis and arterial stiffness, while atherosclerosis and arterial stiffness were positively correlated (Figure 1). This contrasts with the phenotypic comorbidity and cardiovascular-related mortality associated with CWP^13^, suggesting protective variants for CWP may increase atherosclerotic risk. Factor loadings (Figure 2) supported this interpretation, with CWP loading positively and cardiovascular traits negatively on the latent factor, indicating directionally opposing susceptibility.

The common factor GWAS estimated SNP-based heritability at 3.44%, reflecting the proportion of latent factor variance explained by common variants. This lower estimate is expected, reflecting shared, not total, genetic liability.

We identified 53 genome-wide significant SNPs across traits, mapping to 13 genes, including *RNF123, IP6K1,* and *BSN* (Table 3). GO enrichment of 424 suggestive SNPs (Supplementary Table 3) revealed synapse organisation and neurodevelopmental pathway enrichment (Supplementary Figure 2).

FUMA analysis confirmed three independent loci (*RNF123, ATP2C1, COMT)*. MAGMA analysis (see Supplementary Figure 3) further reinforced chromosome 3’s relevance. Tissue-level expression revealed significant DEG enrichment in skeletal muscle (Figure 4) and suggestive enrichment in brain regions, aligning with neurogenic processes and supporting a pain-driven latent factor.

*HYAL1*, *HYAL2*, and *HYAL3* emerged in the post-GWAS analysis. GOMF and KEGG pathway enrichment analysis (Figure 5) highlighted glycan-related immune functions, including hyalurononglucosaminidase activity and glycosaminoglycan degradation, implicating ECM remodelling and immune signalling as central mechanisms underlying the latent factor. Chromatin interaction and eQTL mapping (Figure 7) linked lead variants to these processes.

While we initially hypothesised that broad immune mechanisms would explain the comorbidity, the results instead suggest that neurogenic and glycan-related mechanisms jointly contribute to the shared genetic liability. Enrichment in neuronal differentiation supports a neurodevelopmental interpretation of the latent factor, with relevance to pain perception and vascular regulation, while signals in glycan metabolism and ECM remodelling suggest immune-related mechanisms linking CWP and cardiovascular traits. ECM remodelling is a key driver of vascular inflammation following injury^22^. These findings implicate CNS function and ECM dynamics in the shared genetic liability.

These results build on Naeini et al.^12^ by explaining and accounting for the shared latent factor and characterising its biological nature. While the latent factor appears to align more closely with nervous system processes, the molecular complexity reveals a broader interplay of neurogenic and immune involvement in CWP-cardiovascular comorbidity.

Our analysis included approximately 5.95 million SNPs (see Supplementary Table 1). We observed heritability estimates of 33.3% for CWP, 17.9% for atherosclerosis, and 3.6% for arterial stiffness (Table 1), closely aligning with published estimates of 33%^10^, 15.5%^23^, and 6.1%^24^, respectively. These converging estimates derived from well-powered studies, strong Z-scores, and low SE values, support the reliability of the genetic correlation estimates and factor model.

The common factor model revealed a single latent factor capturing shared genetic liability across CWP, atherosclerosis, and arterial stiffness (Figure 2). Genetic correlations (Figure 1) revealed inverse relationships between CWP and both cardiovascular traits, with a positive correlation observed between atherosclerosis and arterial stiffness. This directional divergence was confirmed by the model, with CWP aligning positively and the cardiovascular traits aligning negatively with the latent factor.

This antagonistic pattern is evident at the SNP level. For instance, *rs1491985* (*RNF123*) exhibited a negative effect size in Rahman et al.^9^ for CWP, indicating a protective effect, but showed a positive effect size for the latent factor, suggesting increased cardiovascular risk. Conversely, *rs10490825* (*ATP2C1*) had a positive effect size for CWP in Rahman et al.^9^ but displayed a negative effect size for the latent factor, indicating reduced cardiovascular susceptibility. Similarly, *rs165599* (*COMT*) had a near-zero effect size in Rahman et al.^9^ but a negative effect for the latent factor, suggesting protective effects against cardiovascular traits. This consistent bidirectional relationship between CWP and two distinct cardiovascular phenotypes, measured using binary and continuous approaches, strengthens the interpretation that the latent factor captures unified SNPs with directionally opposing effects.

This reflects antagonistic pleiotropy, where the same genetic variants have opposing effects across traits. While CWP and cardiovascular comorbidity is well documented^11^, these results reveal a more complex relationship., with some SNPs increasing CWP liability while reducing cardiovascular risk, and others showing the reverse.

Directionally opposing effects have been reported in evolutionary studies of coronary artery disease (CAD), a clinical manifestation of atherosclerosis. CAD-associated SNPs have been linked with increased reproductive success^25^, suggesting positive selection of these variants despite greater cardiovascular risk in later life. The consistency across genetic correlations and factor loadings in our study supports a model where variants increasing cardiovascular risk may reduce CWP susceptibility. This may reflect an evolutionary trade-off, with SNPs conferring late-life cardiovascular risk persisting due to early-life protection against CWP.

While effective for capturing shared genetic liability, the single-factor model may oversimplify genetic relationships. SNPs with shared effects between some, but not all traits, may be overlooked, potentially excluding some biologically relevant SNPs and altering pathway representation.

Despite these constraints, several genome-wide significant SNPs stood out, offering insight into the biological processes shaping the shared genetic liability.

Genome-wide significant SNPs passing Q P-value filtering mapped to 13 unique genes (Table 3), including *RNF123, IP6K1*, and *BSN*. These loci showed homogenous effects across CWP, atherosclerosis, and arterial stiffness, aligning with the latent factor model. *RNF123* has been implicated in CWP studies^9^, supporting its relevance to the latent factor.

Post-GWAS analysis using FUMA^21^, identified 95 suggestive SNPs and confirmed three independent genomic risk loci: *rs1491985* (*RNF123*), *rs10490825* (*ATP2C1*), and *rs165599* (*COMT*) (see Supplementary Table 4). Chromatin interaction and eQTL mapping data (Figure 7) supported these loci, and their convergence across SNP-level, gene-level, and functional mapping reinforces their key contribution to the latent factor.

*RNF123* encodes an E3 ubiquitin ligase involved in protein degradation and cell cycle regulation, protective against CWP^9^. It also has E3-independent functions, regulating antiviral immune response through RIG-I-like receptor signalling^26^, suggesting broader relevance in immune modulation.

*ATP2C1* encodes *SPCA1*, a Golgi apparatus calcium and manganese ion transporter maintaining intracellular homeostasis^27^. Calcium signalling disruptions are linked with neuronal dysfunction^28^ and maladaptive pain processing^29^. Rare *ATP2C1* loss-of-function mutations cause Hailey-Hailey disease, a blistering skin disorder. The condition has shown responsiveness to low-dose naltrexone^30^, a treatment also trialled in fibromyalgia^31^, suggesting overlapping calcium-mediated pain mechanisms.

*COMT* has been extensively linked to CWP through its role in catecholamine metabolism. Low gene activity is associated with greater pain sensitivity^32^ and increased vulnerability to fibromyalgia^33^, migraines^34^, and temporomandibular disorder^35^. *rs4680* has been associated with pain sensitivity modulation in specific chronic pain conditions^36^, although replication across large-scale GWAS has been inconsistent. The lead variant identified in our analysis, r*s165599,* is a 3’ UTR variant independent of *rs4680.* Although it did not replicate in Rahman et al.^9^, who concluded that any role of *COMT* in CWP is likely modest, its emergence as a lead gene in our analysis suggests that *COMT* may still contribute to the shared genetic liability between CWP and cardiovascular traits.

Post-GWAS analysis revealed HYAL gene family enrichment in glycan degradation and hyaluronidase activity (Figure 5).

Tissue-specific expression patterns of the HYAL genes provided additional nsight (Figure 6). *HYAL2* was most expressed in adipose and arterial tissues, with additional activity across the brain and skeletal muscle. *HYAL1* showed moderate adipose expression, and *HYAL3* was enriched in some brain regions. These patterns support glycan-degrading enzymes in ECM and vascular dynamics^22^ within the shared genetic architecture, implicating vascular remodelling as a mechanism linking CWP and cardiovascular traits.

GOMF analysis identified enrichment in hyalurononglucosaminidase and hexosaminidase activity, while KEGG analysis revealed glycosaminoglycan degradation (Figure 5). These findings, centred around *HYAL1*, *HYAL2*, and *HYAL3,* were identified through chromatin interaction and eQTL mapping (Figure 7).

Glycosaminoglycan degradation, driven by hyaluronidase activity, is central to ECM remodelling, influencing immune signalling, vascular repair, and tissue structure^37^. Hyaluronidase activity can influence inflammatory cytokines, support tissue regeneration, and accelerate wound healing by stimulating granulation and angiogenesis^38^. Its recurrence across multiple layers of FUMA analysis highlights these processes as key mediators of the latent factor linking CWP, atherosclerosis, and arterial stiffness.

GO enrichment also revealed an overrepresentation of neurodevelopmental pathways, including synapse organisation, neuron development, and cell junction assembly (see Supplementary figure 2), suggesting CNS connectivity and communication involvement in the shared genetic liability. The convergence of neurodevelopmental and ECM-related pathways implies that the latent factor does not represent a single biological system, but instead emerges from the intersection of immune, structural, and neurovascular mechanisms, supporting a complex, multisystem model unifying pain, vascular, and inflammatory biology.

Identifying a predominantly pain-driven latent factor has clinical implications. While individuals with CWP experience increased cardiovascular morbidity phenotypically^11^, our findings suggest that variants protective against CWP may increase cardiovascular susceptibility. Although behavioural or environmental factors may elevate cardiovascular risk in CWP, the latent factor reflects genetic protection against CWP at the cost of cardiovascular susceptibility. This challenges the view of unified risk between CWP and cardiovascular traits, supporting antagonistic pleiotropy and highlights the need to disentangle phenotypic comorbidity from genetic liability in complex disease research.

The intersection of glycan degradation, ECM remodelling, and neurodevelopmental pathways highlights therapeutic opportunities targeting ECM-related processes in CWP. Post-GWAS identification of glycan-degrading enzymes suggests that modulating hyaluronan activity or restoring ECM homeostasis may reduce atherosclerosis in CWP. Recent evidence implicating altered HYAL2 methylation in blood as a predictive biomarker for preclinical coronary heart disease and stroke further reinforces its clinical relevance in vascular–pain comorbidity^39^.

There are several limitations to the study. The single-factor model may have oversimplified complex genetic patterns, potentially missing SNPs shared between only two traits. Additionally, the custom per SNP loop created to resolve matrix singularity errors and exclude problematic SNPs may have introduced bias.

Secondly, all GWAS summary statistics were obtained from UK Biobank. The reliance on a single sample and lack of replication in independent datasets limits robustness. Finally, GWAS data based solely on European ancestry, may limit generalisability to diverse populations^40^.

Future research should replicate these findings in independent, diverse cohorts to validate the latent factor beyond European populations. Functional characterisation of *RNF123*, *ATP2C1*, *COMT*, and the *HYAL* family using CRISPR to knockout or modulate expression could confirm their roles in ECM remodelling and glycan degradation in pain and vascular disease models. Longitudinal phenotyping could clarify how shared liability influences the onset and severity of atherosclerotic outcomes in CWP individuals. Unannotated SNPs should be prioritised to avoid missing meaningful signals. While Naeini et al.^12^ identified a single latent factor, a multi-factor GenomicSEM approach may capture overlapping, but distinct pathways, capturing trait-specific effects or SNPs influencing subsets of phenotypes masked by the single-factor design.

These directions offer potential in refining our understanding of the genetic relationship between CWP, atherosclerosis, and arterial stiffness. By highlighting antagonistic pleiotropy and the potential of ECM-targeted interventions in CWP to reduce atherosclerosis, this study reframes comorbidity as a product of shared but opposing genetic influences, challenging conventional assumptions and offering a new lens on complex disease comorbidity.

## Methods

### Data Sources

This study used publicly available GWAS summary statistics from Rahman et al.^9^, which identified genetic loci associated with CWP. The dataset included 7,778,994 SNPs from 6,914 CWP cases and 242,929 controls from UK Biobank.

Unlike the ACR 1990 criteria, requiring pain to persist for at least three months in all body quadrants and the axial skeleton (Wolfe et al., 1990), this study adopted a CWP case definition using self-reported data directly from the UK Biobank phenotype file. Participants were asked “*In the last month have you experienced any of the following that interfered with your usual activities?*” (Data Field 6159). Possible answers included “pain all over the body”, pain at specific regions of the body (head, face, neck/shoulder, back, stomach/abdomen, hip, knee), or “none of the above”.

CWP cases included participants reporting “pain all over the body” lasting more than 3 months (Data Field 2956, 3404, 3414, 3571, 3741, 3773, 3799, 4067), simultaneous pain lasting more than 3 months in the knee, shoulder, hip, and back, or fibromyalgia diagnoses (Data Field 20002), including symptom descriptions that led to clinical diagnosis.

Controls included those reporting no pain, “pain all over the body” lasting less than 3 months, or only non-musculoskeletal pain (head, face, stomach/abdomen) lasting more than 3 months. To minimise the risk of case-control misclassification, individuals with self-reported diagnoses of rheumatoid arthritis, polymyalgia rheumatica, arthritis not otherwise specified, systemic lupus erythematosus, ankylosing spondylitis, and myopathy were excluded based on the 20002 data field.

To investigate shared genetic risk factors between CWP and atherosclerosis, publicly available GWAS summary statistics for coronary atherosclerosis (I9_CORATHER) were incorporated into the analysis. This dataset, provided by Neale Lab^41^, comprised 13,586,588 SNPs from 14,334 cases and 346,860 controls of European ancestry, with genotyping conducted using HRC-imputed arrays^42^.

Coronary atherosclerosis in the UK Biobank was identified using hospital inpatient records, based on the International Classification of Diseases, 10^th^ Revision (ICD-10). Participants with a primary or secondary I25.1 diagnosis (Atherosclerotic Heart Disease), were selected from data field 41270, which records hospital admission diagnoses, ensuring all cases were clinically confirmed. The condition is characterised by atherosclerotic plaque accumulation in the coronary arteries, causing narrowing and ischemic complications.

To broaden the analysis of shared genetic architecture, publicly available GWAS summary statistics for arterial stiffness were integrated into the analysis. This dataset from UK Biobank comprised 9,851,867 SNPs from 151,053 individuals of European ancestry, providing information on SNPs associated with arterial stiffness, a key marker for cardiovascular risk.

Arterial stiffness was assessed using the measurement of pulse wave velocity using the finger volume waveform (Data Field 21021). The time taken for the pulse wave to travel through the arterial tree and reflect back in the finger was used to calculate the stiffness index. This was determined by dividing the individual’s height by the time between waveform peaks. As participant height data was unavailable during the initial assessments, the stiffness index was calculated after the arterial stiffness probe had been used, once height information became available. For this analysis, data field 11971, containing the derived pulse wave velocity arterial stiffness index, was used. This dataset was generated using the GWAS pipeline and Pheasant-derived variables, based on the output from data field 21021^41^.

Shared risk variants were identified using GenomicSEM (v0.0.5) in the Linux-based high-performance computing environment CREATE^43^. A Conda-managed R (4.2) environment was used to ensure package compatibility. A common factor model was conducted to capture the genetics of the shared latent factor contributing to the pathology of CWP, atherosclerosis, and arterial stiffness, and give potential insight into their shared biological networks. This modelling approach builds on Grotzinger et al.^44^ who demonstrated its effectiveness in explaining shared genetic influences in common complex conditions.

### Summary Statistics Processing and Quality Control

GWAS summary statistics files for CWP, atherosclerosis, and arterial stiffness were pre-processed for compatibility with GenomicSEM. QC filtering was applied using GenomicSEM munge() function. As part of this process, we applied the HapMap3 SNP reference panel which retained 1,033,180 SNPs from the CWP dataset, 1,200,792 SNPs from the atherosclerosis dataset, and 1,202,434 SNPs from the arterial stiffness dataset for covariance matrix calculation. This panel captured the most relevant genetic signals while excluding low-quality variants that could introduce statistical noise.

### LDSC and Covariance Matrix Construction

Following the initial QC, the processed summary statistics were analysed using LDSC with the precomputed 1000 Genomes European LDSC reference panel to calculate the genetic covariance structure. LDSC merged summary statistics files with the reference panel, removing non-matching SNPs and preserving genomic alignment across traits. Additionally, LDSC applied chi-squared (χ²) filtering to exclude outlier SNPs; however, in this analysis, all retained SNPs fell within the trait-specific thresholds specified by the model.

LDSC produced heritability (h²) estimates based on SNPs and estimated genetic correlations between traits. For binary traits, heritability was converted to the liability scale, while for the continuous arterial stiffness, heritability was estimated based on the observed scale. This process also generated the genetic covariance matrix, representing the genetic overlap between CWP, atherosclerosis, and arterial stiffness, and the sampling covariance matrix, which accounts for the statistical uncertainty and sampling variability in GWAS studies. The LDSC step does not retain individual SNP associations but instead summarises the genome-wide genetic relationship between traits and serves as input for the common factor model.

### Preparing Common Factor GWAS and Ancestry Alignment

For common factor GWAS, we processed the original GWAS summary statistics files to create a unified file containing SNP information for CWP, atherosclerosis, and arterial stiffness. This ensured that SNPs and effect sizes (β) remained consistent across traits. β and SE values were scaled to unit-variance, enabling direct SNP comparison across traits.

To ensure ancestry alignment, we filtered for European-ancestry variants from the 1000 Genomes Phase 3 reference panel, which was originally obtained as individual chromosome files and then merged into a unified reference panel using bcftools. Variants with a minor allele frequency less than 0.01 were excluded to reduce statistical noise and reduce the likelihood of false positive associations. Following filtering, 5,952,753 SNPs were used to generate a combined summary statistics file.

### GenomicSEM Common Factor Model

LDSC matrix and the SNP-level genetic associations were combined to estimate SNP effects on the latent factor underlying CWP, atherosclerosis, and arterial stiffness. Retained SNPs were reintegrated with the LDSC-derived covariance matrices to model polygenic influence and estimate SNP associations across traits, accounting for genetic pleiotropy and sampling variability.

The model specified a single latent factor, with CWP, atherosclerosis, and arterial stiffness as the observed traits. The factor loading for CWP was fixed at 1.0 to define the scale of the latent variable, while loadings for atherosclerosis and arterial stiffness were estimated from the summary statistics. This structure allowed for the evaluation of whether the latent factor aligned more closely with pain or cardiovascular traits, providing initial insights into risk directionality.

A conceptual overview of the model used in the common factor GWAS, illustrating how the LDSC-derived covariance matrices and harmonised summary statistics data are used to estimate individual SNP effects on the latent factor is shown in Supplementary Figure 3.

In several chromosomes, complete matrix singularity errors were triggered during model estimation. To address this, a custom per SNP loop was created using R’s tryCatch() function, excluding and logging problematic SNPs while allowing the remaining analysis to proceed. This approach removed a small subset of SNPs, ensuring numerical stability and preserving results where model estimation was successful.

### Post-GWAS Analyses

Q-Q and Manhattan plots were generated using all SNPs from the common factor GWAS output. This approach enabled the detection of polygenic enrichment and genome-wide signal distribution. Genes were mapped from SNPs with suggestive association (*P* < 5e-6) and retained for GO enrichment analysis, based on evidence of polygenicity across traits and the likely contribution of small-effect variants to shared pathways. The output from common factor GWAS was uploaded to FUMA for functional annotation, enabling interpretation of the results through eQTL, chromatin interactions, and tissue-specific expression analysis.

By integrating CWP, atherosclerosis, and arterial stiffness GWAS summary data, we aimed to identify shared genetic risk loci and better understand the potential molecular or regulatory pathways linking chronic pain to atherosclerosis and arterial stiffness.

## Data Availability

GWAS summary statistics for chronic widespread pain (CWP) are publicly available via Zenodo at https://zenodo.org/records/4459546. GWAS summary statistics for coronary atherosclerosis (I9_CORATHER) are available from the IEU OpenGWAS Project at https://gwas.mrcieu.ac.uk/datasets/ukb-d-I9_CORATHER/. GWAS summary statistics for arterial stiffness (UK Biobank field 11971) are available from the IEU OpenGWAS Project at https://gwas.mrcieu.ac.uk/datasets/ukb-b-11971/. Individual-level UK Biobank data are available through application at https://www.ukbiobank.ac.uk.

The HapMap3 SNP list used for quality control and reference alignment is available at https://alkesgroup.broadinstitute.org/LDSCORE/w_hm3.snplist.bz2. The 1000 Genomes Phase 3 reference panel used for imputation and ancestry alignment can be accessed at https://mathgen.stats.ox.ac.uk/impute/1000GP_Phase3.html.

The FUMA platform used for post-GWAS annotation and functional mapping is available at https://fuma.ctglab.nl/.

## Code Availability

Analyses were performed using the GenomicSEM package (Grotzinger et al., 2019), available at https://github.com/MichelNivard/GenomicSEM. Custom scripts for data preprocessing, LDSC pipeline management, and per-SNP GenomicSEM model execution are available upon reasonable request to the corresponding author.

## Funding acknowledgements

### Funding

This work was supported by Disc4All, CORE Wellcome grant WT 212904/Z/18/Z, and Versus Arthritis grant 22467.

## Acknowledgements

Acknowledgements – This study was conducted by Dúalta McGrath as part of his undergraduate research under the supervision of Maryam Kazemi Naeini and Professor Frances MK Williams. The results presented in this manuscript have not been previously presented elsewhere.

