## Supplementary Information for "Common factor GWAS identifies shared risk loci between chronic widespread pain, atherosclerosis and arterial stiffness"

GWAS Dataset Summary and SNP Filtering

Supplementary Table 1: Overview of GWAS Datasets and SNP Retention across Common Factor GWAS Analytical Steps

| Trait | Sample size (Total / Cases / Controls) | SNPs in original GWAS | SNPs used for LDSC (HapMap3 QC) | SNPs in Common Factor GWAS |
|---|---|---|---|---|
| CWP | 249,843 / 6,914 / 242,929 | 7,778,994 | 1,033,180 | 5,952,753 |
| Atherosclerosis | 361,194 / 14,334 / 346,860 | 13,586,588 | 1,200,792 | 5,952,753 |
| Arterial Stiffness | 151,053 / - / - | 9,851,867 | 1,202,434 | 5,952,753 |

*Supplementary table 1 legend: Sample sizes and SNP counts in each trait dataset. For CWP and atherosclerosis, sample sizes include counts of cases and controls. Arterial stiffness was measured as a continuous trait. Columns report SNP counts in the original GWAS summary statistics, SNPs retained after HapMap3 filtering, and the final SNP set included in the common factor GWAS. All values were recorded after each relevant data processing stage.*

SNP heritability estimates.

SNP-based heritability ($h^2$) was estimated for each trait using Linkage Disequilibrium Score Regression (LDSC). The heritability estimates for CWP, atherosclerosis, and arterial stiffness were consistent with the polygenic nature of these traits. All traits produced highly significant P-values, suggesting that SNP heritability estimates were unlikely to result from sampling variability. The low SE values further support the precision and reliability of these estimates.

These results indicate that while all three traits are polygenic, the proportion of variance explained by common SNPs is substantially higher for CWP and atherosclerosis than for arterial stiffness.

Supplementary Table 2: SNP Heritability Estimates

| Trait | $h^2$ | SE | P-value |
|---|---|---|---|
| CWP | 0.3326 | 0.0284 | 1.2745e-31 |

| | | | |
|---|---|---|---|
| Atherosclerosis | 0.1789 | 0.0161 | 1.2544e-28 |
| Arterial Stiffness | 0.0362 | 0.0040 | 1.3045e-19 |

*Supplementary table 2 legend: SNP heritability ($h^2$) estimates for each trait, calculated using LDSC. Values were derived from HapMap3-filtered SNPs. For binary traits (CWP and atherosclerosis) estimates were converted to the liability scale. Continuous traits (arterial stiffness) were estimated based on the observed scale. SE values reflect the uncertainty in each estimate. P-values represent the significance of heritability estimates.*

SNP Association Results

Supplementary Table 3: Summary of SNPs with Suggestive and Genome-Wide Significant Associations

| Significance Threshold | Total SNPs Identified |
|---|---|
| P < 5e-6 (Suggestive) | 590 |
| P < 5e-6 (Suggestive) and Q P > 0.05 | 424 |
| P < 5e-8 (Genome-wide) | 78 |
| P < 5e-8 (Genome-wide) and Q P > 0.05 | 53 |

*Supplementary table 3 legend: Summary of SNPs showing suggestive and genome-wide association with the latent factor following the common factor GWAS. SNPs with P < 5e-6 and Q P > 0.05 were retained for biological pathway analysis. Genome-wide significant SNPs (P < 5e-8) that passed Q P-value filtering were manually extracted.*

Supplementary Figure 1: Q-Q Plot of SNPs in the Common Factor GWAS Output

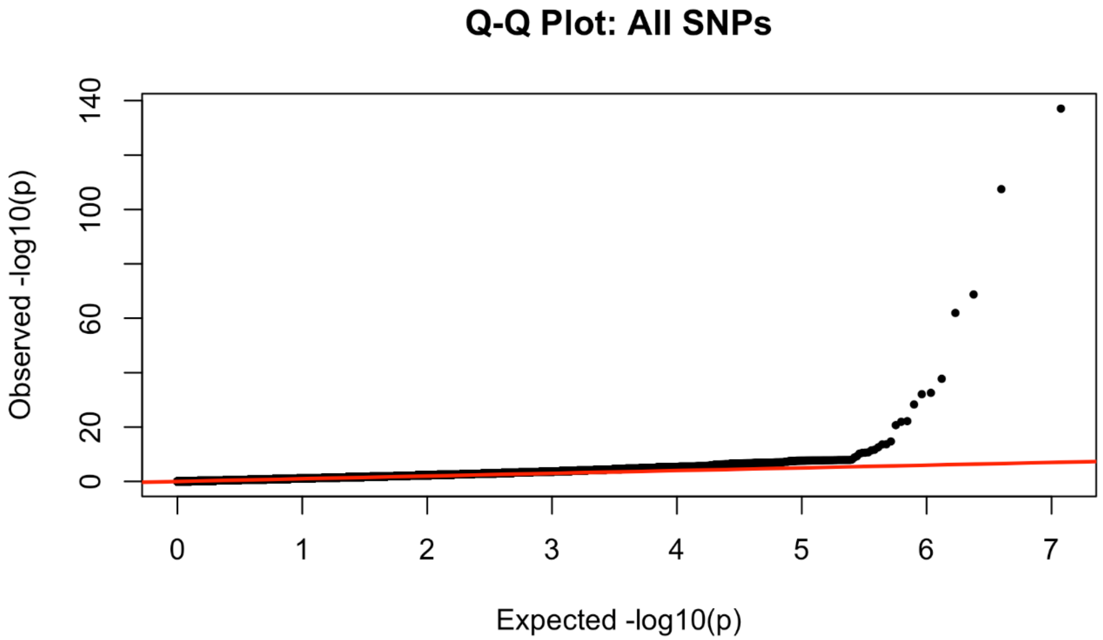


*Supplementary figure 1 legend: Q-Q plot of SNP associations from the common factor GWAS. Each point represents the $-\log_{10}(P)$ value of a SNP plotted against the expected value under the null hypothesis of no association with the latent factor. The red diagonal line represents the null distribution. Most SNPs align closely with the null line, indicating appropriate model calibration. The upward deviation of the tail suggests an excess of SNPs with lower P-values and may reflect polygenic effects, population stratification, or both.*

Enriched Neurodevelopmental Processes

GO enrichment analysis was performed using g:Profiler on genes mapped from 424 SNPs that exceeded suggestive significance (P < 5e-6) and passed Q P-value filtering (Q P > 0.05) to investigate potential biological processes underlying the shared genetic liability. Enriched terms, shown in Figure 3, were predominantly neurodevelopmental, including synapse organisation, neuron development, and nervous system development. Additional enrichment was observed in cellular structure and signalling pathways such as cell junction organisation.

Supplementary Figure 2: Enriched Gene Ontology Biological Processes among SNPs Associated with the Latent Factor

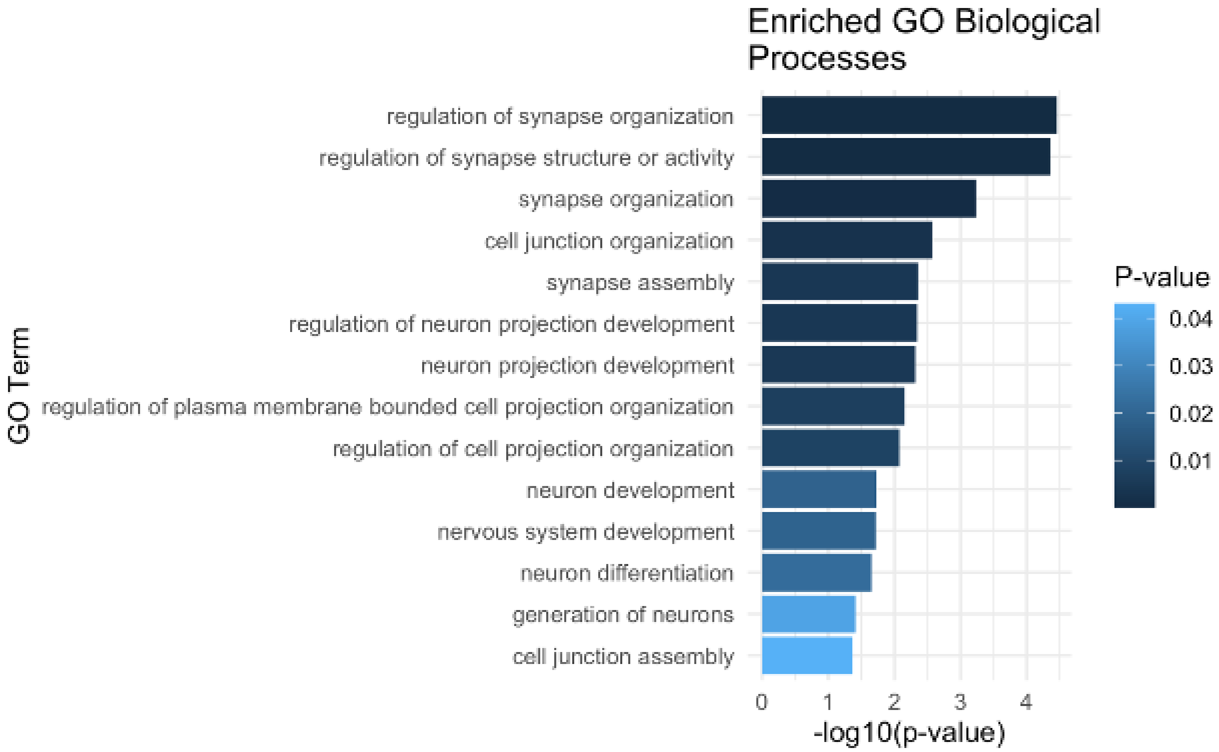


*Supplementary figure 2 legend: Bar plot of the top enriched GO Biological Processes underlying the latent factor based on genes mapped from 424 SNPs that exceeded suggestive significance (P < 5e-6) and passed Q P-value filtering (Q P > 0.05), SNPs were mapped to genes using the g:Profiler package, and enrichment was assessed using the default gSCS multiple testing correction. Bars are ordered by $-log_{10}(P)$ values, with darker blue bars indicating stronger enrichment.*

### Genomic Risk Loci and Gene-Based Analysis

Supplementary Table 4: Independent Genomic Risk Loci identified by FUMA from the Common Factor GWAS

| SNP | CHR | BP | P-value | Gene |
|---|---|---|---|---|
| rs1491985 | 3 | 49739507 | 1.1906E-08 | RNF123 |
| rs10490825 | 3 | 130696383 | 1.2879E-08 | ATP2C1 |
| rs165599 | 22 | 19956781 | 1.3208E-08 | COMT |

*Supplementary table 4 legend: Genomic risk loci identified by FUMA based on lead SNPs passing genome-wide significance (P < 5e-8) and linkage disequilibrium (LD) structure. Gene annotations were assigned using the Open Targets platform by selecting the highest variant-to-gene score for each locus. These loci represent distinct genomic regions contributing to the latent factor, with decimal values rounded to four decimal places.*

Supplementary Figure 3: Gene-based Manhattan Plot of Common Factor GWAS using FUMA

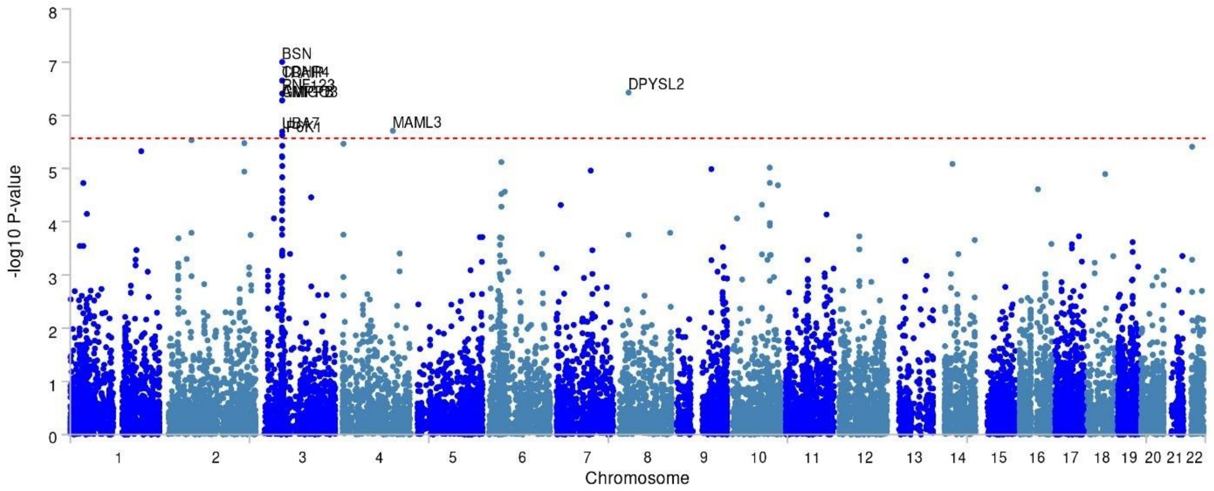


*Supplementary figure 3 legend: The plot displays gene-level association results from MAGMA, using the SNP results from common factor GWAS. 18,530 protein-coding genes were tested, with genome-wide significance set at P < 2.698e-6 (Bonferroni correction). Each point on the plot represents a gene, and the genome-wide significance threshold is indicated by the red horizontal line.*

Analytical Model

Supplementary Figure 4: Conceptual Model of GenomicSEM Common Factor GWAS

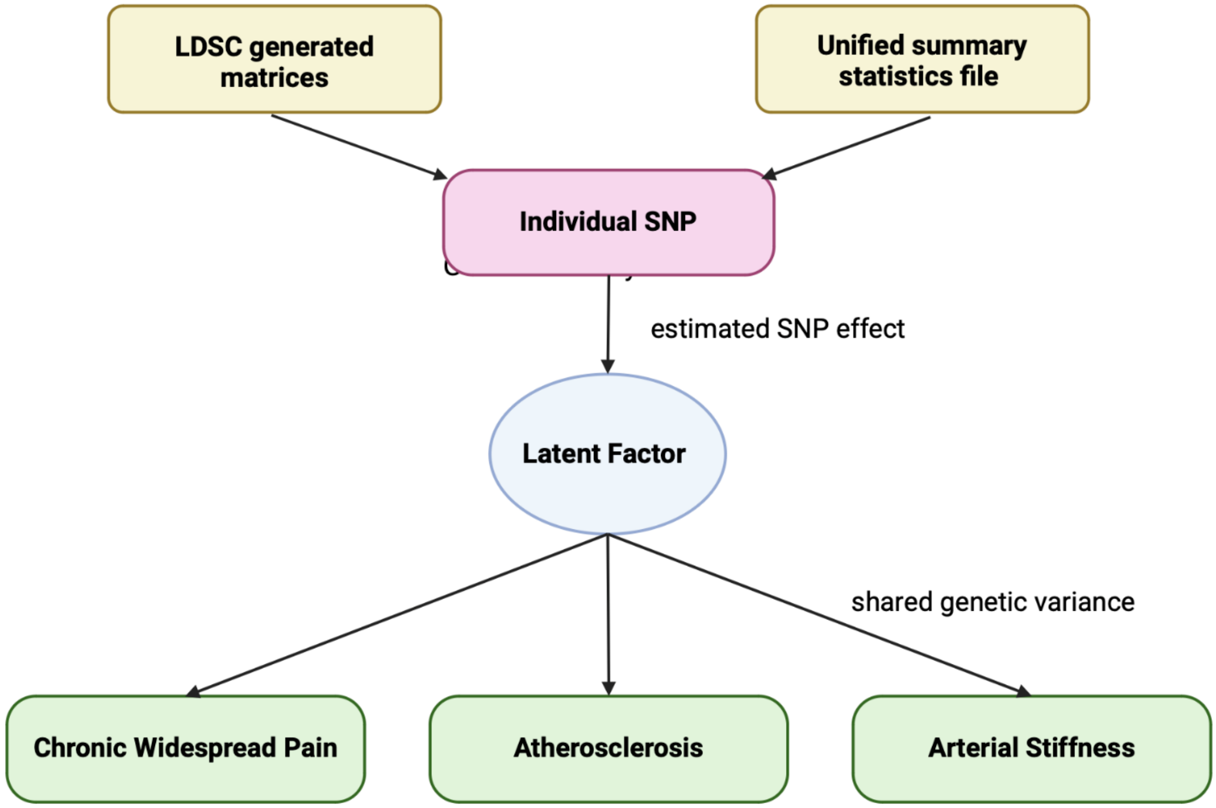


*Supplementary figure 4 legend: Conceptual model of the common factor GWAS. Harmonised SNP-level summary statistics across all traits combined with LDSC-derived genetic covariance and sampling covariance matrices are used to estimate individual SNP associations with the latent factor. The latent factor represents the shared genetic risk liability in CWP, atherosclerosis, and arterial stiffness. This figure was created using Biorender (n.d.).*

Supplementary References:

1. Biorender. BioRender. *Biorender* at <https://biorender.com>
